# Large language models for automatable real-world performance monitoring of diagnostic decision support systems: a comparison to manual doctor panel review in a prospective clinical study

**DOI:** 10.1101/2025.09.20.25336227

**Authors:** Fabienne Cotte, Marcel Schmude, Philipp Bode, Oula Suliman, Filipa Dias Lourenço, Miguel Paiva Pereira, Nisha Kini, Vera Hartenstein, Allesandro Muscoloni, Lisa Stroux, Victor Hertz, Sebastian Köhler, Valerio Morelli, Henry Hoffmann, Peter Engerer, Stephen Gilbert, Kirsten Gray, Tauseef Mehrali, Micaela Seemann Monteiro, Pedro Flores

## Abstract

**Background:** Diagnostic decision support systems (DDSS) are increasingly deployed at scale, yet their diagnostic accuracy is insufficiently monitored once integrated into care. Traditional post-market surveillance relies on clinician review, which is costly, slow, and difficult to sustain. Large language models (LLMs) may offer a scalable and potentially automatable solution, but their performance in real-world monitoring remains unknown.

**Methods:** We conducted a diagnostic accuracy substudy within ESSENCE, a prospective evaluation of Ada Health’s DDSS integrated into Portugal’s largest private healthcare network. Clinical notes and ICD-10 diagnoses from 498 encounters were anonymised and classified using a filter–map–match framework. Manual clinician review served as the reference standard. We compared eligibility classification and condition mapping between clinicians and GPT-4.1 and GPT-5, and assessed diagnostic accuracy of two DDSS versions using both reference sets.

**Findings:** Manual review classified 385 of 498 encounters (77·3%) as eligible for diagnostic comparison. GPT-5 reproduced these classifications with 84·7% accuracy (κ=0·57), showing high sensitivity but only moderate specificity. Among 347 encounters judged eligible by both approaches, GPT-5 exactly matched clinician-assigned diagnoses in 93·6% and proposed clinically plausible alternatives in 3·5%. Diagnostic accuracy estimates based on manual versus GPT-5 mappings were statistically indistinguishable at Top-1 and Top-3 across the full analyzable sets, with one significant difference at Top-5. In the overlapping 346 cases, no statistical differences were observed. Across both reference sets, the experimental DDSS version outperformed the original only at the Top-5 threshold.

**Interpretation:** LLMs can reproduce clinician review of real-world diagnostic encounters with close agreement. While GPT-5 performed comparably to clinicians for condition mapping, the eligibility filtering step - deciding which encounters should enter the diagnostic-accuracy analysis - remains the main source of divergence and is the priority for improvement. Embedding such approaches into health systems could enable automated and continuous performance and safety monitoring and support regulatory compliance. Broader evaluations across diverse care settings are needed to establish generalisability and equity impact.

**Funding:** German Federal Ministry of Education and Research (NextGenerationEU, PATH project).

## Background

Artificial intelligence (AI) has moved from proof-of-concept studies to approved tools in routine care across nearly all domains of medicine. AI now interprets radiology, guides ultrasound, supports pathology, triages symptoms, predicts deterioration, and optimises operations(1–5). Every stage of care delivery is being re-examined through the lens of AI(6).

Robust evidence is needed to validate benefits, ensure safe use, and meet regulatory requirements for demonstrations of performance and clinical value, including methods in the postmarket phase, i.e., post-market surveillance and post-market studies (known as post-market clinical follow-up)(7,8). Yet most evaluations rely on curated test sets, reader studies, or single-centre trials(9). These establish feasibility but rarely capture the heterogeneity of practice, where performance can deteriorate once exposed to diverse patients and workflows(10–12). Silent-mode trials help bridge this gap by embedding AI in clinical settings without influencing care, exposing biases and operational challenges before full deployment. In Kenya, an LLM tool for error detection was run in shadow mode to uncover workflow issues in primary care(4). In the UK, a machine-learning framework for predicting hospital-acquired infections in patients with neurological impairments underwent a year-long silent-mode evaluation, demonstrating superior risk stratification compared with early warning scores(13).

Pre-deployment quality control is also essential. Asgari et al (2025) describe a clinician-in-the-loop framework (CREOLA) that stress-tests documentation assistants with a medical-device-style risk scheme, permitting only versions that meet safety thresholds to progress(14).

Once implemented, AI generates continuous real-world data - an opportunity and obligation to monitor performance, safety, and equity at scale(15). Surveillance should be embedded in continuous improvement cycles using process control, A/B testing, and governance-linked monitoring, yet few organisations have operationalised such systems(7). Traditional post-market evaluation relies on clinician review of AI outputs(16): reliable but costly, poorly scalable and almost impossible to automate. Alternatives link AI predictions to registry-confirmed diagnoses, readmissions, or mortality(12,17), or track whether clinicians accept or reject AI outputs (“passive labelling”)(7,11,18). Patient- and clinician-reported outcomes highlight usability and trust but remain subjective and intermittent(7).

An emerging strategy is “AI to monitor AI.” LLMs have been tested for automated chart review and adverse drug reaction detection(19,20). These systems replicate tasks previously reliant on clinicians, offering scale, consistency and automatability at lower cost. Yet they are not equivalent to humans: LLMs excel at structured extraction but struggle with ambiguity or missing data, risking error amplification. Human reviewers, by contrast, interpret context and detect what is absent(19). The most promising models therefore combine both, with LLMs surfacing structured outputs and clinicians adjudicating uncertain or high-risk cases.

A key domain where this approach could be transformative is monitoring diagnostic decision support systems (DDSS). For systems generating condition suggestions, a central challenge is assessing alignment with confirmed diagnoses(21). Until now, this has relied on panels of physicians judging concordance with clinical diagnoses(16). The task is further complicated by the nature of clinical documentation: recorded diagnoses are often ambiguous or coded at a high level of generality, may reflect only part of a multimorbid presentation, or remain provisional while investigations are ongoing.

### Objectives

This study aimed to (1) develop a reproducible methodology for assessing DDSS outputs against confirmed diagnoses using a framework that accounts for diagnostic ambiguity and context, and (2) evaluate whether LLMs can replicate or scale this process by automating diagnostic-context classification and condition mapping with sufficient reliability to support post-market surveillance.

## Methods

### Study Design and Setting

This diagnostic substudy was nested within ESSENCE, a prospective quality-improvement evaluation of a digital diagnostic decision support system (DDSS) developed by Ada Health (Berlin, Germany) and implemented in Portugal’s largest private healthcare network (CUF). Adults (≥18 years) who completed a Portuguese-language symptom assessment in the myCUF mobile application between Nov 1, 2023, and Oct 31, 2024, provided electronic consent, and shared their report with a CUF clinician were eligible. For this substudy, we included all encounters with both an ICD-10–coded diagnosis and clinical notes available in the electronic health record (EHR).

### Data Collection Procedures

Data collection procedures are outlined in Figure 1. The full ESSENCE protocol has been reported previously (Cotte et al 2025, preprint). In brief, participants completed a symptom assessment and pre- and post-assessment care intentions, and shared the report with CUF physicians as part of routine care. Healthcare-seeking behavior was tracked through EHR review and follow-up email surveys. A research assistant extracted unstructured clinical notes, ICD-10-coded diagnoses, and study-related metadata into a secure electronic system.

**Figure 1.**
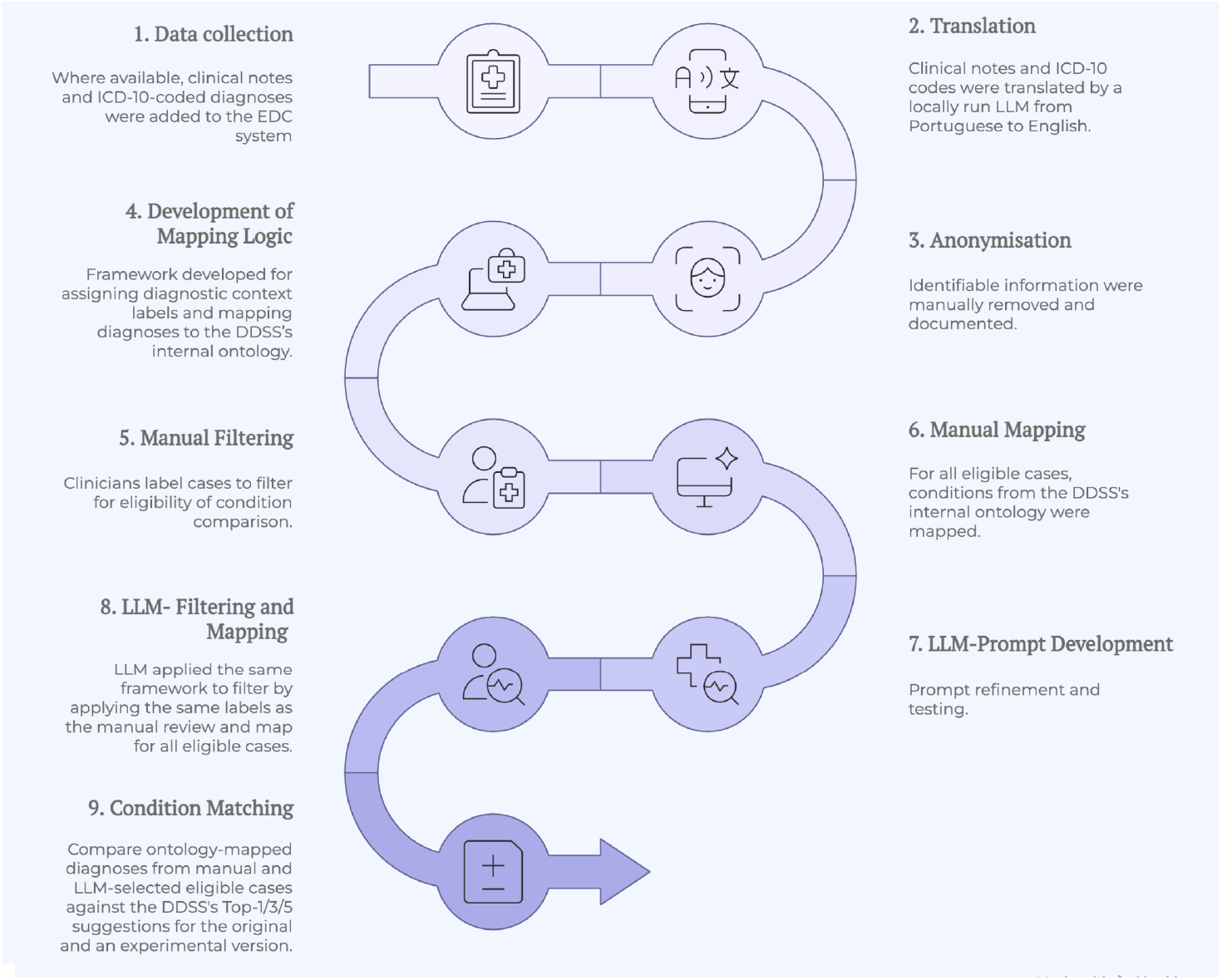
Data collection procedures.

All free-text notes and diagnoses were translated from Portuguese to English using a locally run Mistral-Nemo-Instruct-2407, following manual anonymisation and approval by the data protection officer. Anonymisation followed standardized rules: removal of names, dates, occupations, and location identifiers, and conversion of exact ages into bands. A summary of all actions and affected cases is provided in Supplementary Table 1.

### Manual filtering and mapping (reference set)

Clinicians familiar with the DDSS ontology reviewed each case to establish a reference standard. They determined whether the clinic encounter addressed the same problem as the DDSS assessment and whether a final diagnosis had been reached. Cases with a different complaint, only a symptom recorded, or unspecific ICD-10 codes without clarifying details were excluded. Definitive diagnoses were mapped to one or more DDSS conditions; broad codes were refined using consultation notes. If no equivalent existed, cases were recorded as “condition not covered.” Reviews were performed independently, with disagreements resolved by consensus and a final reviewer ensuring consistency. Definitions of all diagnostic-context labels are provided in Supplementary Table 2.

### LLM-based filtering and mapping

We next evaluated whether a LLM could reproduce this workflow. The model received the same anonymised and translated notes, ICD-10 codes, and user-entered symptoms from the DDSS assessment. Prompts instructed the LLM to act as a medical coding specialist and toB follow the same four-step logic: (1) filtering cases based on the match between the user’s complaint and the clinical encounter, (2) assessing the specificity of the final diagnosis, (3) refining unspecific diagnoses using details from the clinical notes, and (4) mapping the final, specified diagnosis to the DDSS’s condition ontology. Few-shot examples of each diagnostic-context label were included to standardise reasoning, and outputs were returned in structured JSON format.

Because the full DDSS ontology exceeded the model’s context window, we created vector embeddings for each DDSS condition (BAAI/bge-large-en-v1.5). For each case, the ICD-10 description and note text were used to query this index, and the top 100 most relevant conditions were inserted into the prompt as the candidate set. To ensure reliability, the model’s temperature was fixed at 0, and ten outputs were generated per case. Final labels and mappings were determined by majority vote. The process was fully automated in Python (OpenAI API).

### Condition Matching

Two case sets were created: one from manual review and one from the LLM. These overlapped but were not identical, reflecting differences in eligibility and mappings. Each set was compared with condition suggestions from two DDSS versions. The original system, based on Ada’s probabilistic reasoning, was the version deployed during ESSENCE (Miller et al. 2020). The experimental version was a compound AI system under development utilising an LLM from the Llama 3 family. It was included in line with principles of post-market surveillance, where monitoring should establish whether newer versions provide measurable improvements. For fair comparison, it was restricted to the same user inputs without generating extra questions.

### Data Analysis

Analyses were conducted in Python (v3.12) and Google Sheets. Diagnostic-context labelling was assessed by overall agreement and Cohen’s κ, with precision, recall, and F1 reported per label. Systematic differences between manual and LLM distributions were tested using χ^2^. Eligibility for diagnostic-accuracy analysis was assessed as a binary outcome, with accuracy, sensitivity, specificity, precision, and F1 reported.

For condition mapping, LLM-assigned diagnoses were compared with clinician mappings. Outputs were classified as exact match (LLM selected one of the clinician-assigned conditions), plausible additional (clinically valid but overlooked by clinicians), or mismatch (clinically inappropriate). Comparative analyses were performed for GPT-4.1 (gpt-4.1-2025-04-14) and GPT-5 (gpt-5-2025-08-07, reasoning effort “high”).

Finally, the diagnostic accuracy of the DDSS was assessed by comparing reference diagnoses - derived from manual clinician review or GPT-5 mapping - with the ranked condition suggestions generated by the system. Accuracy was evaluated at the Top-1, Top-3, and Top-5 levels, defined as the proportion of cases in which the reference diagnosis appeared among the first one, three, or five system outputs. Diagnostic accuracy was reported with 95% CIs. Two-sample tests for proportions were used for unpaired comparisons, and McNemar’s test was applied for paired analyses. Comparisons between original and experimental DDSS versions were conducted within each reference set using paired methods. Cases in which the real condition was not modelled by the DDSS were still included in the diagnostic accuracy analysis.

### Missing Data Handling

Encounters without ICD-10–coded diagnoses or consultation notes were excluded, as these data formed the reference standard and could not be imputed. Reasons for exclusion are listed in Supplementary Table 3.

### Ethics and Data Governance

The ESSENCE study received ethical approval from the Comissão de Ética para a Investigação Clínica (2204JJ351, 2309JJ660) and was registered on ClinicalTrials.gov (NCT06846957). All participants provided electronic informed consent. Data were anonymised and stored securely in Teamscope, and only de-identified records were used for LLM analysis. Procedures complied with the Declaration of Helsinki and ISO 14155:2020.

### Funding

The Federal Ministry of Education and Research (Germany) funded the study but had no role in design, data collection, analysis, interpretation, or writing.

## Findings

### Participant characteristics and inclusions

A total of 512 participants were included. The mean age was 39·2 years (SD 12·4; median 37 years [IQR 30–47]); 80·3% were aged 18–49 years. Females accounted for 57·6% (295/512). The mean number of presenting complaints (PCs) was 2·21 (SD 1·43), higher in females (2·40) than males (1·95; p=0·00036). The most common specialties were otorhinolaryngology/ENT (24·0%), orthopedics/trauma surgery (18·2%), gynecology (14·6%), and gastroenterology (13·5%). Details are provided in Table 1. Of 1,470 participants in the main cohort, confirmed healthcare-seeking behavior was available for 721 (49·0%). Within this group, 512 cases had both an ICD-10 code and consultation notes. Fourteen illustrative cases were set aside as examples for prompt development (Supplementary Table 4), leaving 498 cases for which labels were assigned by both manual review and the LLM (Figure 2).

**Table 1.**
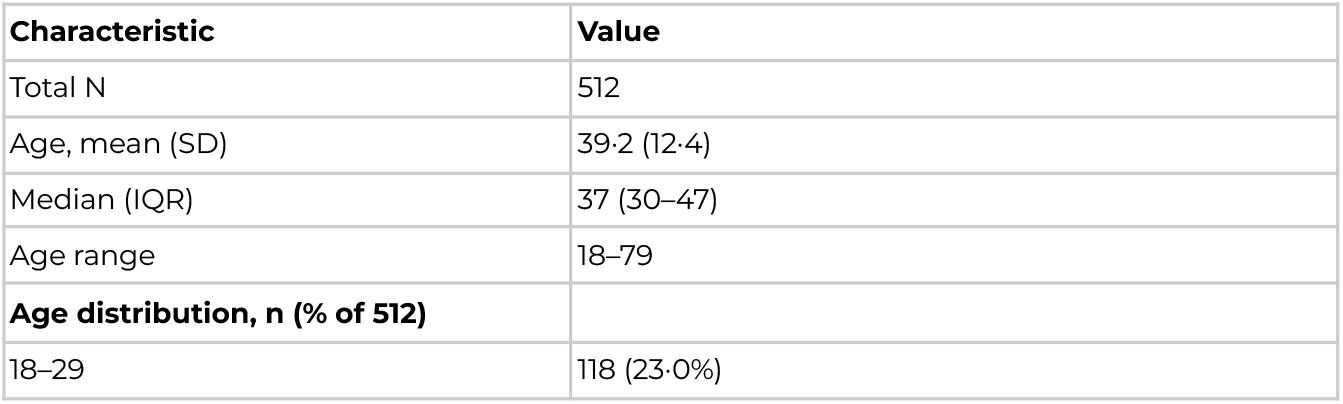
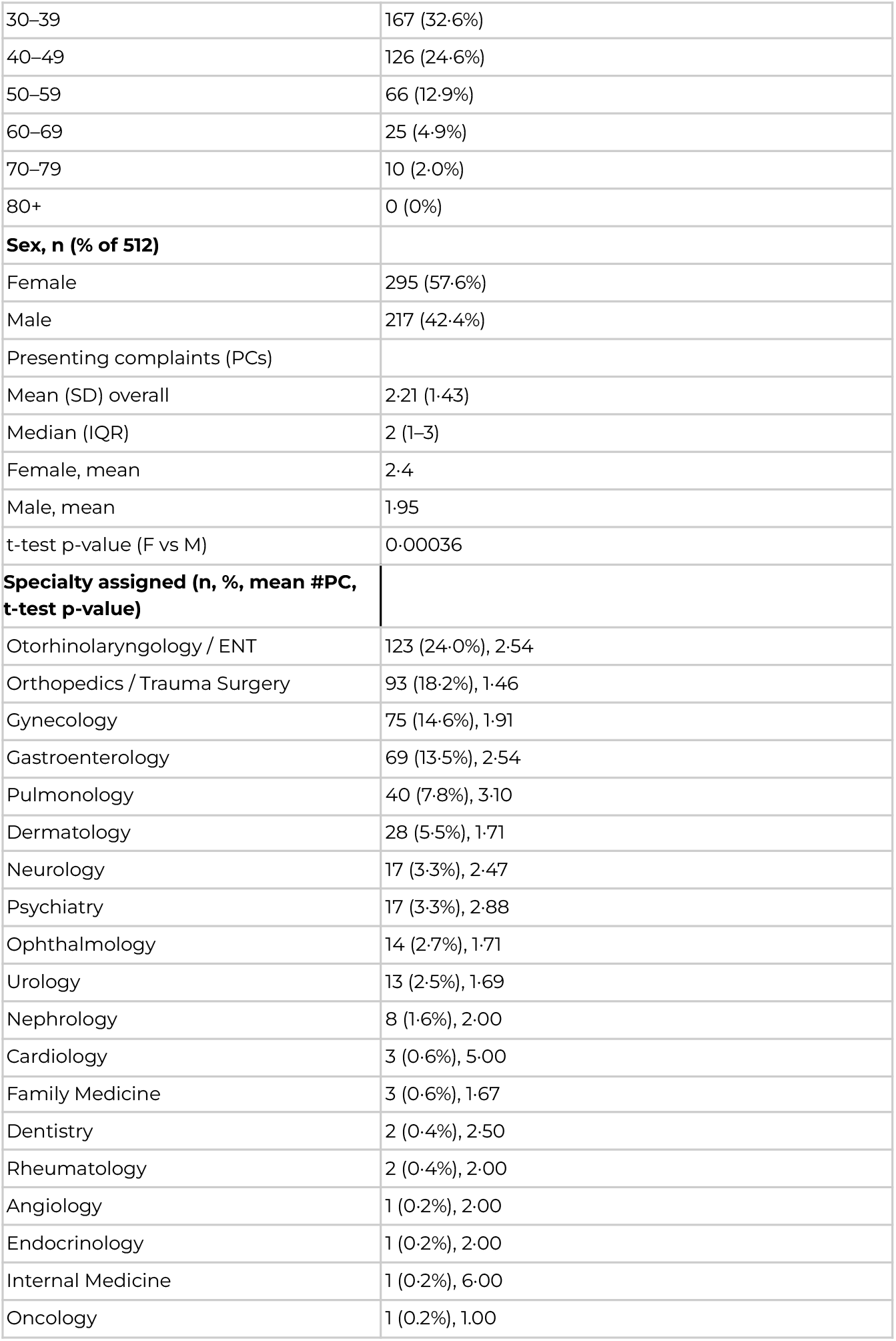
Participant characteristics

**Figure 2.**
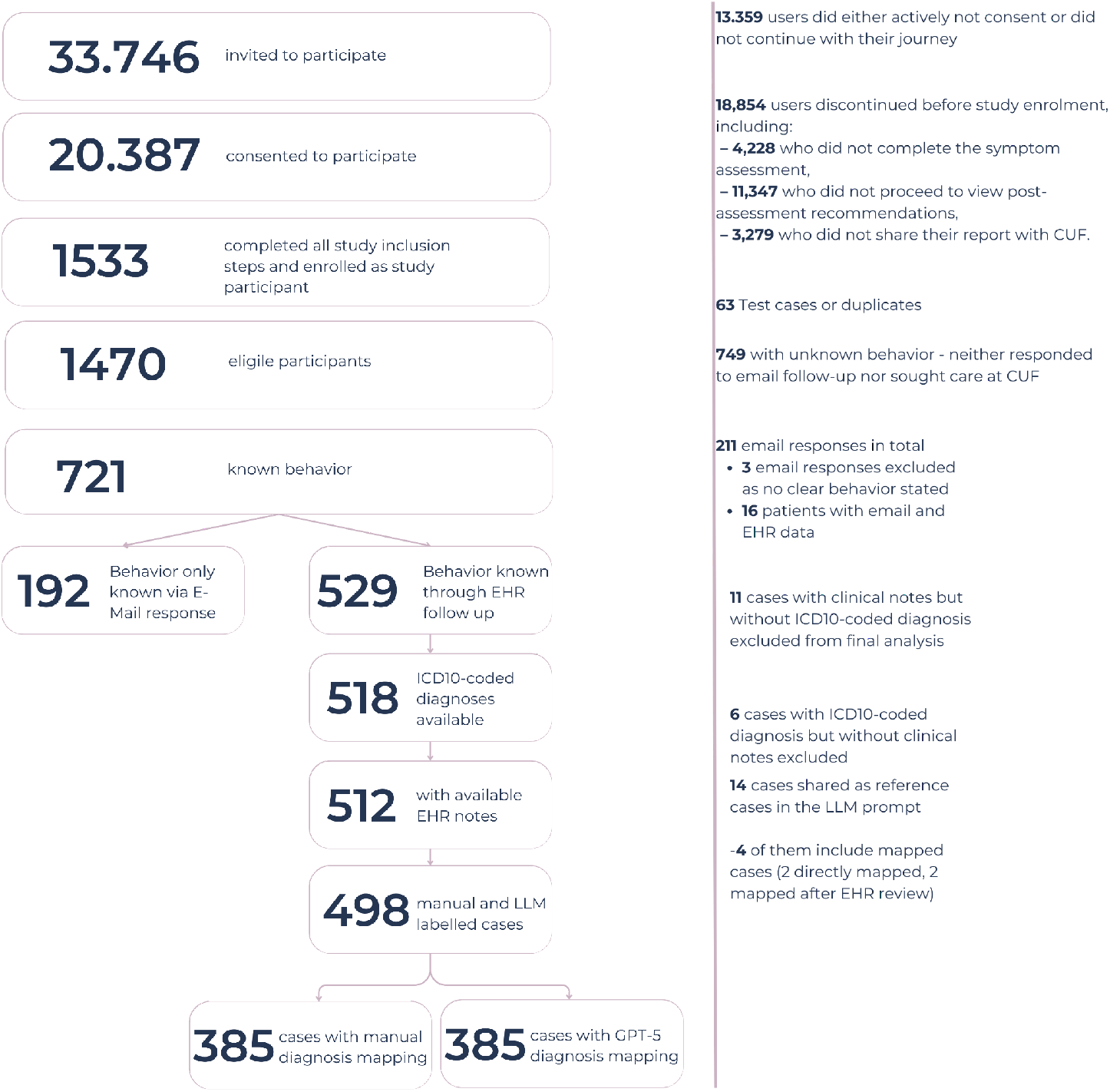
STROBE Inclusion flowchart.

### Anonymisation Outcomes

All free-text notes were anonymised before LLM mapping. Of 512 notes, 235 (45·9%) required modification, most commonly conversion of exact ages to bands (n=186) and removal of dates (n=59), occupations (n=35), or locations (n=32). Names were removed in 18 cases. Over half of notes (283/512, 55·3%) required no changes (Supplementary Table 1).

### Filtering and Mapping

Manual review classified 385/498 encounters (77·3%) as eligible and distributed the remaining 113 (22·7%) across exclusion categories: 46 (9·2%) “no diagnosis–symptom,” 12 (2·4%) “no diagnosis–unspecific code,” 25 (5·0%) “multimorbidity,” 21 (4·2%) “different complaint,” and 9 (1·8%) “not-covered condition” (Table 2).

**Table 2.** Diagnostic-context label distribution and per-label performance

| Label | Manual n (%) | GPT-5 n (%) | Both | Missed | Precision | Recall | F1 |
| --- | --- | --- | --- | --- | --- | --- | --- |
| **Mappable** | 385 (77.3) | 385 (77.3) | 347 | 38 | 0.9 | 0.9 | 0.9 |
| **No Dx – Symptom** | 46 (9.2) | 41 (8.2) | 30 | 16 | 0.73 | 0.65 | 0.69 |
| **No Dx – Unspecific code** | 12 (2.4) | 27 (5.4) | 5 | 7 | 0.19 | 0.42 | 0.26 |
| **Multimorbidity** | 25 (5.0) | 11 (2.2) | 10 | 15 | 0.91 | 0.4 | 0.56 |
| **Different complaint** | 21 (4.2) | 22 (4.4) | 11 | 10 | 0.5 | 0.52 | 0.51 |
| **Not-covered condition** | 9 (1.8) | 12 (2.4) | 6 | 3 | 0.5 | 0.67 | 0.57 |
| **Total** | 498 (100) | 498 (100) | — | — | — | — | — |

GPT-5 reproduced this overall distribution, also assigning 385/498 (77·3%) encounters as eligible. Among these, 347/385 (90·1%) overlapped with manual review. The model missed 38/385 (9·9%) manually eligible cases, while conversely assigning diagnoses in 38/113 (33·6%) cases that clinicians had excluded.

At the binary level (eligible/ineligible), GPT-5 achieved 84·7% accuracy (95% CI 81·3–87·6), with sensitivity 90·1% (95% CI 86·7–92·7) and specificity 66·4% (95% CI 57·3–74·4). Agreement beyond chance was moderate (κ=0·57, 95% CI 0·50–0·64) (Table 3). Performance was robust for eligible cases (precision 0·90, recall 0·90, F1=0·90) but less consistent for exclusion categories, where unspecific codes were frequently over-assigned and multimorbidity under-recognised.

**Table 3.** Performance of GPT-4.1 and GPT-5.0 for eligibility classification and condition mapping (n=498)

|  | GPT-4.1 | GPT-5.0 |
| --- | --- | --- |
| **Eligibility classification** |  |  |
| True Positives (TP) | 374 | 347 |
| False Negatives (FN) | 11 | 38 |
| False Positives (FP) | 71 | 38 |
| True Negatives (TN) | 42 | 75 |
| Manual eligible cases | 385 | 385 |
| Model eligible cases | 445 | 385 |
| Accuracy (TP+TN / N) | 83.5% | 84.7% |
| Sensitivity (recall for eligible) | 97.1% | 90.1% |
| Specificity (recall for ineligible) | 37.2% | 66.4% |
| Precision (PPV for eligible) | 84.0% | 90.1% |
| F1 score (for eligible) | 90.1% | 90.1% |
| Cohen's $\kappa$ (95% CI) | 0.42 (0.33–0.52) | 0.57 (0.50–0.64) |
| **Condition mapping** |  |  |
| Exact match | 354/374 (94.7%) | 324/347 (93.6%) |
| Should be considered (plausible addition) | 8/374 (2.1%) | 12/347 (3.5%) |
| No overlap (mismatch) | 12/374 (3.2%) | 10/347 (2.9%) |
| No output | – | 1/347 (0.3%) |

### Model comparison: GPT-4.1 vs GPT-5.0

Both models reproduced manual eligibility classifications with high overall accuracy (Table 3). GPT-4.1 achieved higher sensitivity (97·1% vs 90·1%) but much lower specificity (37·2% vs 66·4%). GPT-5·0 provided a more balanced profile with higher precision (90·1% vs 84·0%) and stronger agreement (κ=0·57 vs 0·42). Running this workflow required $5.08 for a single pass with GPT-4.1 and $44.40 for GPT-5.

### Condition mapping performance

Among jointly eligible cases, GPT-5 exactly matched one of the clinician-assigned diagnoses in 324/347 (93·6%) and proposed an additional clinically plausible condition in 12 (3·5%). Disagreement occurred in 10 cases (2·9%), and no output in one (0·3%). GPT-4.1 showed a similar profile: 354/374 (94·7%) exact matches, 8 (2·1%) plausible additions, and 12 (3·2%) mismatches. When exact matches and plausible additions were combined, overall alignment was 97·1% for GPT-5 and 96·8% for GPT-4.1. Full distributions are shown in Table 3.

### Consistency

To assess the internal consistency of GPT-5’s label assignment, we generated eight independent completions per case and quantified the distribution of distinct labels. In 385/498 cases (77.3%), all completions converged on the same label. Among the remaining 113 cases, two distinct labels were suggested in 63/498 (12.7%), three in 38/498 (7.6%), and four in 12/498 (2.4%). Even when multiple labels appeared, the majority label typically dominated: for two-label cases, 70% (44/63) showed a 6–2 or 7–1 split.

### Matching comparison

For the diagnostic accuracy analyses, we included all cases, including those where the DDSS did not explicitly model the underlying condition, yielding 394 manual and 397 GPT-5–mapped cases (Table 4).

**Table 4.** Diagnostic accuracy of DDSS using manual and GPT-5 reference mapping.

| Reference set | N | Version | Top-1 % (95% CI) | Top-3 % (95% CI) | Top-5 % (95% CI) |
| --- | --- | --- | --- | --- | --- |
| Manual-mapped | 394 | Original | 47.2 % (42.3–52.1) | 71.6 % (66.5–76.3) | 78.4 % (73.2–83.1) |
|  | 394 | Experimental | 48.0% (47.5–52.9) | 67.5 % (62.4–72.2) | 81.2 % (76.0–85.8) |
| GPT-5-mapped | 397 | Original | 43.1 % (38.3–48.0) | 65.2 % (60.3–69.8) | 71.3 % (66.6–75.6) |
|  | 397 | Experimental | 44.1 % (39.2–49.0) | 67.0 % (62.2–71.5) | 75.1 % (70.6–79.1) |
| Manual-mapped | 346 | Original | 46.8 % (41.6–52.1) | 72.8 % (67.9–77.2) | 80.3 % (75.8–84.2) |
|  | 346 | Experimental | 48.3 % (43.1–53.5) | 68.8 % (63.7–73.4) | 83.5 % (79.3–87.1) |
| GPT-5- | 346 | Original | 48.0 % (42.8–53.2) | 71.4 % (66.4–75.9) | 78.0 % (73.4–82.1) |
| mapped | 346 | Experimental | 49.1 % (43.9–54.4) | 74.0 % (69.1–78.3) | 81.2 % (76.8–85.0) |

With manual mapping, Top-1 accuracy was 47·2% (42·3–52·1) for the original and 48·0% (47·5–52·9) for the experimental DDSS version; Top-3 accuracy was 71·6% (66·5–76·3) versus 67·5% (62·4–72·2); and Top-5 accuracy was 78·4% (73·2–83·1) versus 81·2% (76·0–85·8). With GPT-5 mapping, Top-1 accuracy was 43·1% (38·3–48·0) versus 44·1% (39·2–49·0); Top-3 accuracy was 65·2% (60·3–69·8) versus 67·0% (62·2–71·5); and Top-5 accuracy was 71·3% (66·6–75·6) versus 75·1% (70·6–79·1).

Across the 385 analyzable cases per set, manual and GPT-5 mapping performed similarly (supplementary table 7). Differences ranged from –4 to +7 percentage points, with only one significant finding: manual mapping achieved higher Top-5 accuracy under the original DDSS version (309/385 [80·3%] vs 283/385 [73·5%]; Δ=+6·8 pp, 95% CI +0·8 to +12·7; p=0·026).

After GPT5 did not provide an output in one case, 346 cases overlapped. For these, results were almost identical (supplementary table 8). McNemar tests showed no significant differences at any threshold, with small differences (–5 to +2 pp) and confidence intervals crossing zero, indicating that GPT-5 mapping was statistically equivalent to manual mapping when applied to the same cases.

Paired comparisons of the original and experimental DDSS versions (supplementary table 9) showed no significant differences at the Top-1 or Top-3 thresholds. At Top-5, however, the experimental version was consistently more accurate: manual 309/396 (78·0%) vs 320/396 (80·8%; Δ=+3·1 pp, 95% CI +0·4 to +5·8; p=0·038) and GPT-5 283/397 (71·3%) vs 298/397 (75·1%; Δ=+3·9 pp, 95% CI +1·2 to +6·6; p=0·009).

## Interpretation

### Summary of results

We evaluated whether large language models (LLMs) can automate real-world monitoring of diagnostic accuracy in digital diagnostic decision support systems (DDSS) using a filter–map–match framework. Of 498 cases, both manual and LLM review classified 385 as eligible for diagnostic accuracy analysis. Within this subset, agreement was high: both approaches classified the same 347 cases (90·1%) as eligible. The LLM additionally marked 38 cases as eligible that clinicians had excluded, while excluding 38 that clinicians had included. GPT-5 mapped the same clinician diagnoses in 93·6% of overlapping cases and suggested plausible alternatives in 3·5%. Diagnostic accuracy was statistically indistinguishable between manual and GPT-5 mapping at Top-1 and Top-3, with one difference at Top-5. In overlapping cases, no significant differences remained, confirming equivalence when eligibility filtering is held constant. Across reference sets, both approaches identified that the experimental DDSS was more accurate than the original only at the Top-5 threshold.

### Filtering–mapping–matching

Comparing DDSS outputs with real-world outcomes requires identifying truly comparable encounters. In practice, the digital assessment and clinic visit may cover different complaints, diagnoses may still be under investigation, or multimorbidity can complicate documentation. In our cohort, only 77·3% of encounters were suitable for diagnostic comparison. Analysing all cases together would conflate incomparable scenarios and distort estimates. A structured filter–map–match workflow - (i) filtering to comparable encounters, (ii) mapping confirmed diagnoses to a shared ontology, and (iii) matching mapped diagnoses to DDSS outputs - offers a more rigorous basis for monitoring.

For filtering, the model achieved high sensitivity and precision but only moderate specificity. This asymmetry is acceptable in surveillance: retaining most eligible cases preserves sample size, while extra inclusions can be resolved downstream. Similar frameworks are being explored for case identification in electronic records. Cheligeer et al, for example, applied a multi-stage method to detect hospital-acquired pulmonary embolism from 10,066 inpatients, of whom only 40 (0·4%) had true events. Models achieved sensitivity 87·5–100% and specificity 94·9–98·9%, but positive predictive values remained low (≈7–17%) because of rarity, with F1 scores peaking at 28·1%(22). Their method efficiently ruled out negatives at scale, but most flagged cases still needed manual review.

One key objective of this research was to identify whether automating the entire filter-map-match workflow with AI would distort estimates of diagnostic accuracy. When comparing the full sets of analyzable cases (385 each), GPT-5 mapping produced results that were statistically indistinguishable from manual review at the Top-1 and Top-3 thresholds, with a single significant difference observed at Top-5.

When restricting the analysis to the 347 overlapping cases, where the task was limited to mapping rather than eligibility filtering, no statistical differences were observed at any threshold. This indicates that the mapping step can already be automated with high reliability. The only residual divergence arose from imperfect eligibility filtering, which accounted for the single significant difference in one of the three comparisons. Importantly, both manual and GPT-5 mapping consistently identified that the experimental DDSS version was significantly more accurate at the Top-5 threshold only.

### Methodological considerations

ICD-10 codes were initially considered as the main comparator, but have well-described limitations: agreement is only moderate at chapter level and poor for specific codes, with high inter-rater variability(23–25). Coding complexity and incomplete documentation often yield generic labels(26,27). We therefore combined ICD-10 review with free-text note analysis, which clarified unspecific codes in several cases. LLMs now make it feasible to screen documentation at scale, with high sensitivity and specificity for phenotyping but only moderate positive predictive value and limited causal attribution, requiring clinician oversight(28).

### Strengths

This study has several strengths. It was conducted in a real-world clinical environment within Portugal’s largest private healthcare network, capturing the heterogeneity of routine practice beyond curated test sets. The inclusion of multiple specialties increases the generalisability of findings. A clinician-adjudicated reference standard ensured reproducibility of comparisons. Finally, the filter–map–match framework was explicitly aligned with clinical reality, and the privacy-preserving pipeline systematically anonymised notes, ensuring GDPR compliance. Finally, we also compared downstream diagnostic accuracy across both manual and GPT-5 mapping approaches and between original and experimental DDSS versions. This allowed us to test how the approaches would behave under real-world performance monitoring conditions.

### Limitations

This study has several limitations. Manual review is an imperfect reference, and in some instances the LLM proposed plausible alternatives not documented by clinicians, highlighting the constraints of the comparator. Case review was conducted by clinicians employed by the DDSS and familiar with its ontology, which may have introduced bias. The analysis was a proof-of-concept, one-off evaluation rather than an implemented surveillance system, and cases without a final diagnosis at data collection were excluded, though some might have become eligible later. Performance was also assessed within a single healthcare network, so results may differ in other settings with different documentation practices or diagnostic distributions. Embedding the workflow in live systems will be essential to establish longitudinal performance, feasibility, and clinical impact.

### Implementation in Clinical Practice

Several lessons emerge for real-world use. First, the core decision is binary- whether a case is eligible for diagnostic comparison. Although a multi-label framework was useful for benchmarking, rare categories performed inconsistently and added little operational value. Prioritising this binary decision offers a more pragmatic foundation for implementation, particularly as the mapping step showed no statistically significant differences between manual and GPT-5 review.

Second, diagnosis is often a longitudinal process rather than a single clinical event. Patients may await test results or referrals before a final diagnosis is reached. Systems should therefore re-analyse such cases once definitive information becomes available, converting encounters initially excluded into eligible cases for analysis.

Third, in-workflow one-click feedback controls offer a practical way to capture clinician judgement at the point of care - for example, by accepting a suggested condition or selecting a better match from a type-ahead ontology. Structured reporting in radiology has shown that even a single click can return machine-readable feedback for continual learning (Fuchs 2024). Although clinicians are unlikely to provide feedback for every consultation, such mechanisms are a scalable and efficient addition, delivering targeted corrections without disrupting workflow.

Finally, privacy must be preserved. Advances in locally deployable anonymisation models now achieve near-perfect sensitivity and character-level accuracy for de-identifying clinical text without data leaving the system(29). Integrating such tools could replace manual redaction and enable GDPR-compliant note review at scale.

### Conclusion

This study shows that large language models can automate key steps in monitoring the diagnostic accuracy of decision support systems. GPT-5 produced results that were highly consistent with clinician review, with the few differences explained by eligibility filtering - deciding which real-world cases could be included in the diagnostic accuracy analysis-rather than by the mapping step itself. Both manual and LLM-based approaches led to the same conclusions about the performance improvements from the original to the experimental version of the DDSS, indicating that LLM-based monitoring can be trusted to detect meaningful changes in system performance without continuous manual review.

While these findings establish feasibility, further work is required to embed the workflow in live systems and to extend it to automated continuous monitoring. Focusing on improving eligibility filtering while including re-evaluation of provisional cases, one-click feedback, and local anonymisation will be critical to test scalability and clinical impact. If operationalised, such pipelines could enable continuous, learning health system monitoring of diagnostic AI.

## Research in Context

### Evidence before this study

Monitoring the diagnostic accuracy of digital decision support systems (DDSS) has traditionally relied on manual clinician review. Although accurate, this approach is labour-intensive and difficult to scale. Previous work shows that large language models (LLMs) can extract information from clinical notes and map diagnoses to standard ontologies, but their ability to fully replace clinical review in continuous surveillance of DDSS performance had not been established.

### Added value of this study

This study tested whether an LLM could automate the filter-map-match workflow used to monitor DDSS accuracy in real-world practice. GPT-5 achieved high agreement with manual review, reproducing clinician-adjudicated diagnoses in most cases and generating comparable diagnostic accuracy estimates. Importantly, both manual and LLM-based workflows led to the same conclusions about relative performance of the original versus experimental DDSS versions. Differences between methods arose mainly from eligibility filtering rather than mapping itself.

### Implications of all the available evidence

These findings show that LLMs can reliably replicate the core elements of clinician-led surveillance of DDSS diagnostic accuracy. The mapping step appears nearly ready for practice, whereas eligibility filtering requires further refinement to ensure consistency. If implemented, such an approach could substantially reduce reliance on manual practitioner panels and enable scalable, near real-time monitoring of DDSS in routine care.

## Data Availability

Data collected for the study, including individual participant data and a data dictionary defining each field in the set, will be made available to others after publication upon reasonable request, subject to approval. Requests for access should be made to the study team at Ada Health. After approval, a signed data sharing agreement will be required before data release.

## Contributors

FC and SG were responsible for conceptualisation and methodology. Data collection and investigation were carried out by FDL, MP, MSM, PF, NK, MS, TM, and FC. Data anonymisation was performed by MS. NK, OS, FC, and KG conducted the manual filtering and mapping of cases to establish the clinician-adjudicated reference set. PB, FC, and OS performed formal analysis and visualisation. FC led writing the original draft in collaboration with all co-authors. VHa, AM, LS, VHe, OS, SK, VM, HH, and PE led the development of the experimental version of the app tested in the study and contributed to review of the manuscript. All authors had full access to aggregated data in the study and contributed to the decision to submit the manuscript for publication.

## Acknowledgment

This work was supported by the Federal Ministry of Education and Research (Bundesministerium für Bildung und Forschung) through the European Union-financed NextGenerationEU program under grant number 16KISA100K, project PATH—”Personal Mastery of Health and Wellness Data.”

