## Supplementary Appendix for "Large language models for automatable real-world performance monitoring of diagnostic decision support systems: a comparison to manual doctor panel review in a prospective clinical study"

**Supplementary Table 1**. Anonymisation Actions Applied to EHR Notes

| Element | Anonymisation Action | Count |
|---|---|---|
| Age | Converted exact age to predefined age bands (e.g. “35” → “30–39”) | 186 |
| Dates | Removed or generalised exact dates for exams, symptoms, travel, procedures, and follow-ups; retained year only | 59 |
| Occupations | Removed all occupation details | 35 |
| Locations | Removed names of hospitals, cities, and countries; travel destinations converted to continent level; tourist status kept unless sensitive | 32 |
| Names | Removed all names of patients, clinicians, and relatives | 18 |
| Rare Diseases | Removed conditions and medications deemed potentially identifying due to rarity (e.g. vitiligo, Ehlers-Danlos, paramyloidosis) | 10 |
| Social Information | Removed social information about patient incl. children’s ages, birthdates, educational level psychotherapy mentions, and family disease history when identifiable | 8 |
| Pregnancy Details | Retained pregnancy week (e.g. “35+2” → “35th week”); removed day-specific or conception dates | 7 |
| Weight & Height | Removed when extreme or irrelevant; included only when clinically necessary | 7 |
| Identifiable Physical Characteristics | Removed specific mentions such as tattoos, shoe size, dominant hand | 3 |
| Genetic / Blood Group Data | Removed references to blood type or rhesus group | 2 |
| No action | No anonymisation actions required | 283 |

**Supplementary Table 2**. Diagnostic Context Labels Used for Mapping

| Label | Definition | Included in Accuracy Analysis |
|---|---|---|

| | | |
|---|---|---|
| Mapped | Final diagnosis was specific and directly mappable to DDSS condition(s) | Yes |
| Unspecific code but specification possible with EHR | ICD-10 code was broad, but final condition could be clarified from clinical notes | Yes |
| No final diagnosis reached – symptom | Final diagnosis was a presenting symptom; no underlying cause confirmed | No |
| No final diagnosis reached – unspecific code | Unspecific ICD-10 code used; clinical note indicated need for further diagnostic work-up | No |
| Multimorbidity | Clinical notes mentioned the symptom assessment topic, but the primary diagnosis addressed a different issue | No |
| Different presenting complaint | Clinical consultation addressed a different medical issue than reported in the symptom assessment | No |
| Condition not covered | A specific diagnosis was made, but no corresponding condition existed in the DDSI's condition library | No |

**Supplementary Table 3:** Incomplete Diagnostic Records

| **ESSENCE-ID** | **Case Type** | **Available Data** | **Summary** | **ICD-10** | **Reason** |
|---|---|---|---|---|---|
| ESSENCE-007265 | Excluded | Clinical notes only | 29M, recurrent anal fissure symptoms; suspected infection; follow-up planned | – | Diagnosis not yet recorded at time of review |
| ESSENCE-007619 | Excluded | Clinical notes only | Elderly woman, asymptomatic gyn visit; obesity limited exam | – | Routine check-up; diagnostic work-up pending |
| ESSENCE-034208 | Excluded | Clinical notes only | Neurological symptoms worsening; referred to neurology | – | Evaluation deferred to specialist |
| ESSENCE-037344 | Excluded | Clinical notes only | Teleconsultation for chronic condition follow-up | – | Ongoing care; no new diagnosis assigned |
| ESSENCE-038312 | Excluded | Clinical notes only | Urticaria triggered by environment; improved with avoidance | – | Diagnosis assumed clinically; not coded |

| | | | | | |
|---|---|---|---|---|---|
| ESSENCE-038774 | Excluded | Clinical notes only | 52F with GI symptoms; colonoscopy normal; CT scan requested | – | Diagnostic clarification pending |
| ESSENCE-045095 | Excluded | Clinical notes only | Wide range of GI and systemic symptoms; tests ordered | – | Diagnostic process ongoing |
| ESSENCE-047268 | Excluded | Clinical notes only | Dry nocturnal cough; cardiac work-up normal | – | No diagnosis yet documented |
| ESSENCE-047398 | Excluded | Clinical notes only | Post-cesarean, consulted for infant's urinary issues | – | Not user-focused visit |
| ESSENCE-048203 | Excluded | Clinical notes only | ENT symptoms for 10 days | – | Likely viral; no diagnosis entered |
| ESSENCE-050861 | Excluded | Clinical notes only | Thyroid-related symptoms; labs ordered | – | Diagnosis deferred pending test results |
| ESSENCE-004372 | Excluded | ICD-10 code only | Atypical facial pain | G50.1 | Clinical notes unavailable; structured diagnosis retained |
| ESSENCE-021857 | Excluded | ICD-10 code only | Benign paroxysmal vertigo | H81.1 | Clinical notes unavailable; structured diagnosis retained |
| ESSENCE-032611 | Excluded | ICD-10 code only | Upper respiratory tract infection | J06.9 | Clinical notes unavailable; structured diagnosis retained |
| ESSENCE-037726 | Excluded | ICD-10 code only | Burnout | Z73.0 | Clinical notes unavailable; structured diagnosis retained |
| ESSENCE-047291 | Excluded | ICD-10 code only | Lumbar pain | M54.5 | Clinical notes unavailable; structured diagnosis retained |
| ESSENCE-047861 | Excluded | ICD-10 code only | Acute tonsillitis | J03 | Clinical notes unavailable; structured diagnosis retained |

**Supplementary Table 4.** Example labels

| ESSENCE_ID | Final MappedConditions | Label | Reasoning |
|---|---|---|---|

| | | | |
|---|---|---|---|
| ESSENCE-018018 | [] | "Condition not covered by Ada" | The final diagnosis is Synovial cyst NOS (ICD-10: M71.3), which refers to a benign fluid-filled sac in the joint or tendon lining. While Ada models ganglion cysts, these are considered clinically and anatomically distinct. Because no appropriate equivalent exists in the Ada condition ontology, this case is categorised as "Condition not covered by Ada." |
| ESSENCE-048736 | [] | "Condition not covered by Ada" | The consultation resulted in a final diagnosis of Tarsal tunnel syndrome (ICD-10: G57.5), a nerve compression disorder affecting the tibial nerve. This condition is not currently represented in Ada's internal ontology. Despite clinical specificity, no match is possible, warranting the label "Condition not covered by Ada." |
| ESSENCE-008748 | [] | "Multimorbidity" | The Ada symptom assessment focused on upper respiratory complaints suggestive of a viral infection. The EHR notes describe otitis media as well as rhinorrhoea and nasal congestion, but only the otitis media is reflected in the ICD-10 diagnosis. Because Ada and the EHR each reflect different aspects of the clinical picture, this is considered a multimorbid presentation. |
| ESSENCE-009288 | [] | "Multimorbidity" | The consultation is primarily coded as gastritis, but EHR documentation also discusses abdominal colic and frequent loose stools (3–4 episodes/day), which were the central complaint in the Ada report. Since the final ICD-10 code does not account for these ongoing GI symptoms, but the EHR does, the case reflects multimorbidity—two relevant but separate clinical complaints. |
| ESSENCE-007352 | ["condition:Balanoposthitis"] | "Mapped" | The final diagnosis is Candidal balanoposthitis (ICD-10: B37.4), indicating inflammation of the glans and foreskin due to a fungal infection. While Ada does not differentiate by pathogen, it models Balanoposthitis as a general inflammatory condition that encompasses common causes, including candida. As the underlying diagnosis and mapped Ada condition align clinically, this case is "Mapped." |
| ESSENCE-003837 | ["condition:AcuteUrticaria"] | "Mapped" | The clinical notes describe an acute allergic reaction in a pregnant patient with skin eruptions and itching that started a few days ago and worsened over time. The trigger is suspected to be a topical cream, and the plan includes stopping the cream and reassessing in 2 days.<br>This presentation indicates a short-term, trigger-associated urticaria episode without evidence of daily wheals for more than six weeks, which would be required to classify it as chronic spontaneous or chronic inducible urticaria. The symptoms are likely to resolve with treatment and avoidance of the trigger, aligning best with acute urticaria. |
| ESSENCE-000609 | [] | "Patient presenting for different medical problem" | The symptom assessment concerned headache, yet the consultation dealt exclusively with right elbow pain. No mention of the headache appears in the clinical documentation. The final ICD-10 code reflects only the musculoskeletal issue. Thus, the presenting complaint differs entirely from the issue addressed during the visit. |

| | | | |
|---|---|---|---|
| ESSENCE-006145 | [] | "Patient presenting for different medical problem" | The user's Ada assessment described symptoms consistent with a common cold. However, the EHR consultation addresses chronic right upper quadrant pain and postprandial nausea, linked to gallbladder pathology. The ICD-10 code is K80.2 – Calculus of gallbladder without cholecystitis, and the plan includes surgical referral for possible cholecystectomy.<br>Because the clinical consultation focused entirely on gastrointestinal complaints unrelated to the respiratory symptoms assessed by Ada, this case represents a different medical problem, and is therefore excluded from accuracy analysis. |
| ESSENCE-042363 | [] | "No final diagnosis reached – unspecific code" | The ICD-10 code assigned is N64.9 – Disorder of breast, unspecified, but the clinical notes reveal that no definitive breast pathology was identified. The breast examination was unremarkable, and the primary clinical findings relate to musculoskeletal chest wall tenderness and an inguinal nodule, with further imaging requested.<br>This suggests that no conclusive diagnosis was established at the time of consultation, and the code used does not reflect a specific condition that can be mapped. The case is therefore classified as "No final diagnosis reached – unspecific code". |
| ESSENCE-045596 | [] | "No final diagnosis reached – unspecific code" | The patient presents with two weeks of isolated diarrhoea and is in otherwise excellent health. While Infectious gastroenteritis and colitis, unspecified (A09) was coded, the EHR states that the etiology remains unclear and plans further diagnostics (e.g., abdominal ultrasound). As no definitive diagnosis has been reached yet, and investigations are ongoing, this case is appropriately labelled as "No final diagnosis reached – unspecific code. |
| ESSENCE-037641 | [] | "No final diagnosis reached – symptom" | Although the ICD-10 code H81.10 – Peripheral vertigo was assigned, the cause of vertigo remains unclear, and the clinical notes explicitly state that multiple diagnostic tests and specialist consultations have been ordered (e.g., VNG, Holter monitor, ENT). The patient presents with vertigo, nausea, and vomiting, but no underlying condition has yet been confirmed, and the note reflects ongoing investigation. Therefore, this case is best classified as "No final diagnosis reached – symptom". |
| ESSENCE-037445 | [] | "No final diagnosis reached – symptom" | The ICD-10 code M25.562 – Pain in left knee reflects a symptom, not a definitive diagnosis. The clinical notes describe recent onset of popliteal fossa pain after physical activity, but no underlying condition is identified. Examination and imaging (X-ray) showed no abnormalities, and the patient was treated symptomatically and referred to orthopaedics for further evaluation.<br>As the cause remains unclear and under investigation, this case is best categorised as "No final diagnosis reached – symptom", with no mapping to a specific condition. |
| ESSENCE-051310 | ["condition:ViralGas | "Unspecific code | The ICD-10 code A09.9 – Gastroenteritis and colitis of unspecified origin is unspecific, but the clinical notes allow for further refinement. The patient presented with abdominal pain, fever, nausea, vomiting, and watery diarrhoea |

| | | | |
|---|---|---|---|
| | troenteritis"] | but specification possible with EHR" | of three days' duration, alongside family members with similar symptoms, strongly suggesting a viral aetiology.<br>There is no indication of bacterial testing or findings, and the presentation aligns with self-limited viral gastroenteritis. Therefore, although the original ICD code is non-specific, the EHR allows a confident mapping to "condition:ViralGastroenteritis". |
| ESSENCE-031699 | ["condition:AcuteLumbosacralRadiculopathy", "condition:MusculoskeletalLowerBackPain"] | "Unspecific code but specification possible with EHR" | The assigned ICD-10 code M25.5 – Pain in joint is broad and does not fully reflect the clinical context. However, the EHR notes clearly describe left-sided lower back pain of 10 days' duration, with associated muscle strain/spasm in the gluteus maximus.<br>While the code points to a general joint pain, the location, muscular involvement, and chronicity described in the notes align more closely with musculoskeletal lower back pain, and potentially acute lumbosacral radiculopathy.<br>Although sciatica is not explicitly mentioned or denied, the distribution and symptoms justify mapping to both "condition:MusculoskeletalLowerBackPain" and "condition:AcuteLumbosacralRadiculopathy". |

**Supplementary Table 5.** Confusion matrix of LLM vs. manual labels

| | pred: Mappable | pred: No Dx – Symptom | pred: No Dx – Unspecific | pred: Multimorbidity | pred: Different complaint | pred: Not-covered | Total |
|---|---|---|---|---|---|---|---|
| true: Mappable | 347 | 9 | 6 | 12 | 7 | 4 | 385 |
| true: No Dx – Symptom | 12 | 30 | 2 | 1 | 1 | 0 | 46 |
| true: No Dx – Unspecific | 5 | 0 | 5 | 0 | 2 | 0 | 12 |
| true: Multimorbidity | 11 | 1 | 1 | 10 | 1 | 1 | 25 |
| true: Different complaint | 8 | 1 | 2 | 1 | 11 | 1 | 21 |
| true: Not-covered | 2 | 0 | 0 | 0 | 1 | 6 | 9 |
| Total | 385 | 41 | 16 | 24 | 24 | 12 | 498 |

**Supplementary Table 6.** Example labels

| ESSEN | Final | Label | Reasoning |
|---|---|---|---|

| CE_ID | MappedConditions | | |
|---|---|---|---|
| ESSENCE-018018 | [] | "Condition not covered by Ada" | The final diagnosis is Synovial cyst NOS (ICD-10: M71.3), which refers to a benign fluid-filled sac in the joint or tendon lining. While Ada models ganglion cysts, these are considered clinically and anatomically distinct. Because no appropriate equivalent exists in the Ada condition ontology, this case is categorised as "Condition not covered by Ada." |
| ESSENCE-048736 | [] | "Condition not covered by Ada" | The consultation resulted in a final diagnosis of Tarsal tunnel syndrome (ICD-10: G57.5), a nerve compression disorder affecting the tibial nerve. This condition is not currently represented in Ada's internal ontology. Despite clinical specificity, no match is possible, warranting the label "Condition not covered by Ada." |
| ESSENCE-008748 | [] | "Multimorbidity" | The Ada symptom assessment focused on upper respiratory complaints suggestive of a viral infection. The EHR notes describe otitis media as well as rhinorrhoea and nasal congestion, but only the otitis media is reflected in the ICD-10 diagnosis. Because Ada and the EHR each reflect different aspects of the clinical picture, this is considered a multimorbid presentation. |
| ESSENCE-009288 | [] | "Multimorbidity" | The consultation is primarily coded as gastritis, but EHR documentation also discusses abdominal colic and frequent loose stools (3–4 episodes/day), which were the central complaint in the Ada report. Since the final ICD-10 code does not account for these ongoing GI symptoms, but the EHR does, the case reflects multimorbidity—two relevant but separate clinical complaints. |
| ESSENCE-007352 | ["condition:Balanoposthitis"] | "Mapped" | The final diagnosis is Candidal balanoposthitis (ICD-10: B37.4), indicating inflammation of the glans and foreskin due to a fungal infection. While Ada does not differentiate by pathogen, it models Balanoposthitis as a general inflammatory condition that encompasses common causes, including candida. As the underlying diagnosis and mapped Ada condition align clinically, this case is "Mapped." |
| ESSENCE-003837 | ["condition:AcuteUrticaria"] | "Mapped" | The clinical notes describe an acute allergic reaction in a pregnant patient with skin eruptions and itching that started a few days ago and worsened over time. The trigger is suspected to be a topical cream, and the plan includes stopping the cream and reassessing in 2 days.<br>This presentation indicates a short-term, trigger-associated urticaria episode without evidence of daily wheals for more than six weeks, which would be required to classify it as chronic spontaneous or chronic inducible urticaria. The symptoms are likely to resolve with treatment and avoidance of the trigger, aligning best with acute urticaria. |
| ESSENCE-000609 | [] | "Patient presenting for differe | The symptom assessment concerned headache, yet the consultation dealt exclusively with right elbow pain. No mention of the headache appears in the clinical documentation. The final ICD-10 code reflects only the musculoskeletal issue. Thus, the presenting complaint differs entirely from the issue addressed during the visit. |

| | | | |
|---|---|---|---|
| | | nt medical problem" | |
| ESSENCE-006145 | [] | "Patient presenting for different medical problem" | The user's Ada assessment described symptoms consistent with a common cold. However, the EHR consultation addresses chronic right upper quadrant pain and postprandial nausea, linked to gallbladder pathology. The ICD-10 code is K80.2 – Calculus of gallbladder without cholecystitis, and the plan includes surgical referral for possible cholecystectomy.<br>Because the clinical consultation focused entirely on gastrointestinal complaints unrelated to the respiratory symptoms assessed by Ada, this case represents a different medical problem, and is therefore excluded from accuracy analysis. |
| ESSENCE-042363 | [] | "No final diagnosis reached – unspecific code" | The ICD-10 code assigned is N64.9 – Disorder of breast, unspecified, but the clinical notes reveal that no definitive breast pathology was identified. The breast examination was unremarkable, and the primary clinical findings relate to musculoskeletal chest wall tenderness and an inguinal nodule, with further imaging requested.<br>This suggests that no conclusive diagnosis was established at the time of consultation, and the code used does not reflect a specific condition that can be mapped. The case is therefore classified as "No final diagnosis reached – unspecific code". |
| ESSENCE-045596 | [] | "No final diagnosis reached – unspecific code" | The patient presents with two weeks of isolated diarrhoea and is in otherwise excellent health. While Infectious gastroenteritis and colitis, unspecified (A09) was coded, the EHR states that the etiology remains unclear and plans further diagnostics (e.g., abdominal ultrasound). As no definitive diagnosis has been reached yet, and investigations are ongoing, this case is appropriately labelled as "No final diagnosis reached – unspecific code. |
| ESSENCE-037641 | [] | "No final diagnosis reached – symptom" | Although the ICD-10 code H81.10 – Peripheral vertigo was assigned, the cause of vertigo remains unclear, and the clinical notes explicitly state that multiple diagnostic tests and specialist consultations have been ordered (e.g., VNG, Holter monitor, ENT). The patient presents with vertigo, nausea, and vomiting, but no underlying condition has yet been confirmed, and the note reflects ongoing investigation. Therefore, this case is best classified as "No final diagnosis reached – symptom". |
| ESSENCE-037445 | [] | "No final diagnosis reached – | The ICD-10 code M25.562 – Pain in left knee reflects a symptom, not a definitive diagnosis. The clinical notes describe recent onset of popliteal fossa pain after physical activity, but no underlying condition is identified. Examination and imaging (X-ray) showed no abnormalities, and the patient was treated symptomatically and referred to orthopaedics for further evaluation.<br>As the cause remains unclear and under investigation, this case is best |

| | | | |
|---|---|---|---|
| | | symptom" | categorised as “No final diagnosis reached – symptom”, with no mapping to a specific condition. |
| ESSENCE-051310 | ["condition:ViralGastroenteritis"] | "Unspecific code but specification possible with EHR" | The ICD-10 code A09.9 – Gastroenteritis and colitis of unspecified origin is unspecific, but the clinical notes allow for further refinement. The patient presented with abdominal pain, fever, nausea, vomiting, and watery diarrhoea of three days’ duration, alongside family members with similar symptoms, strongly suggesting a viral aetiology.<br>There is no indication of bacterial testing or findings, and the presentation aligns with self-limited viral gastroenteritis. Therefore, although the original ICD code is non-specific, the EHR allows a confident mapping to "condition:ViralGastroenteritis". |
| ESSENCE-031699 | ["condition:AcuteLumbosacralRadiculopathy", "condition:MusculoskeletalLowerBackPain"] | "Unspecific code but specification possible with EHR" | The assigned ICD-10 code M25.5 – Pain in joint is broad and does not fully reflect the clinical context. However, the EHR notes clearly describe left-sided lower back pain of 10 days' duration, with associated muscle strain/spasm in the gluteus maximus.<br>While the code points to a general joint pain, the location, muscular involvement, and chronicity described in the notes align more closely with musculoskeletal lower back pain, and potentially acute lumbosacral radiculopathy.<br>Although sciatica is not explicitly mentioned or denied, the distribution and symptoms justify mapping to both "condition:MusculoskeletalLowerBackPain" and "condition:AcuteLumbosacralRadiculopathy". |

**Supplementary Table 7.** Diagnostic accuracy of DDSS using manual and GPT-5 mapping (unpaired comparison of full analyzable sets, n=385 each).

| **Version** | **Threshold** | **Manual % (n/N)** | **GPT-5 % (n/N)** | **Δ (pp)** | **95% CI Δ** | **p-value** |
|---|---|---|---|---|---|---|
| Original | Top-1 | 48.3% (186/385) | 44.4% (171/385) | 3.9 | –3.1 to +10.9 | 0.28 |
| | Top-3 | 73.2% (282/385) | 67.3% (259/385) | 6 | –0.5 to +12.4 | 0.07 |
| | Top-5 | 80.3% (309/385) | 73.5% (283/385) | 6.8 | 0.8 to +12.7 | 0.026 |
| Experimental | Top-1 | 49.2% (189/384) | 45.5% (175/385) | 3.8 | –3.3 to +10.8 | 0.3 |
| | Top-3 | 69.1% (266/385) | 69.1% (266/385) | 0 | –6.5 to +6.5 | 1 |
| | Top-5 | 83.3% (320/384) | 77.4% (298/385) | 5.9 | 0.3 to +11.5 | 0.038 |

**Supplementary Table 8**. Diagnostic accuracy of DDSS using manual and GPT-5 mapping in overlapping cases (paired comparison, n=346).

| Version | Threshold | Manual % (n/N) | GPT-5 % (n/N) | Δ (pp) | 95% CI for Δ | McNemar p |
|---|---|---|---|---|---|---|
| Original | Top-1 | 46.8% (162/346) | 47.9% (166/346) | –1.2 | –7.0 to +4.6 | 0.68 |
| | Top-3 | 72.8% (252/346) | 71.4% (247/346) | 1.4 | –4.8 to +7.6 | 0.71 |
| | Top-5 | 80.3% (278/346) | 78.0% (270/346) | 2.3 | –3.0 to +7.6 | 0.42 |
| Experimental | Top-1 | 48.3% (167/346) | 49.1% (170/346) | –0.9 | –6.9 to +5.1 | 0.8 |
| | Top-3 | 68.8% (238/346) | 73.9% (256/346) | –5.1 | –11.1 to +0.9 | 0.1 |
| | Top-5 | 83.5% (289/346) | 81.2% (281/346) | 2.3 | –2.9 to +7.6 | 0.42 |

**Supplementary Table 9.** Statistical comparison of original vs. experimental DDSS versions (paired McNemar tests, n≈385 per reference set)

| Reference set | Threshold | Original % | Experimental % | Δ (pp) | 95 % CI for Δ | McNemar p |
|---|---|---|---|---|---|---|
| **Manual** | **Top-1** | **47.1** | 47.7 | 1 | –1.1 to +3.2 | 0.48 |
| | Top-3 | 71.3 | 67.2 | –4.2 | –10.2 to +1.9 | 0.21 |
| | Top-5 | 78.1 | 80.8 | 3.1 | 0.4 to +5.8 | 0.038 |
| **GPT-5** | Top-1 | 43.1 | 44.1 | 1 | –1.2 to +3.3 | 0.5 |
| | Top-3 | 65.2 | 67 | 1.8 | –1.1 to +4.7 | 0.3 |
| | Top-5 | 71.3 | 75.1 | 3.9 | 1.2 to +6.6 | 0.009 |
